# Genome-wide association study of ischemic stroke risk in Sickle Cell confirms *ADAMTS2, CDK18*, uncovers 12 novel loci

**DOI:** 10.1101/2022.08.22.22279082

**Authors:** Eric Jay Earley, Shannon Kelly, Fang Fang, Cecília Salete Alencar, Daniela de Oliveira Werneck Rodrigues, Dahra Teles Soares Cruz, Ester Sabino, Brian Custer, Carla Dinardo, Grier P. Page, the International Component of the NHLBI Recipient Epidemiology and Donor Evaluation Study (REDS-III) and the NHLBI Trans-Omics for Precision Medicine (TOPMed) Consortium

## Abstract

**Background:** Ischemic stroke is a common complication of sickle cell disease (SCD) and without screening or intervention can affect 11% of children with SCD before the age of 20. This study sought to find genetic biomarkers for risk of stroke occurring at younger ages.

**Methods:** Within the Trans-Omics for Precision Medicine (TOPMed), a genome-wide association study (GWAS) of ischemic stroke was performed on 1,333 individuals with SCD from Brazil (178 cases, 1155 controls). Via a novel proportional hazards analysis approach, we searched for variants associated with strokes occurring at younger ages.

**Results:** Fourteen genomic regions were associated with early ischemic stroke at genome wide significance (P<5×10^−8^). This included variants near two genes which have been previously linked to non-SCD early onset stroke (<65 years): *ADAMTS2* (rs147625068, P= 3.70 × 10^−9^) and *CDK18* (rs12144136, P= 2.38 × 10^−9^), respectively. Individuals harboring multiple risk alleles exhibited increasing rates of stroke at earlier timepoints (P < 0.001, Gehan-Wilcoxon) than those carrying only one. Enrichment tests suggest systemic dysregulation of gene expression in the hypothalamus (*P* = 0.03, FDR), substantia nigra (*P* = 0.03), spleen (*P* = 0.005) and coronary (*P* = 0.0005), tibial (*P* = 0.03) and aorta arteries (*P* = 0.03.

**Conclusions:** This findings from this study support a model of shared genetic architecture underlying ischemic stroke risk between SCD individuals and non-SCD individuals <65 years. In addition, results suggest an additive liability due to carrying multiple risk alleles.

## INTRODUCTION

Ischemic stroke is one of the most common comorbidities of sickle cell disease (SCD), occurring 20 times more frequently than seen in non-SCD children^1,2^. Without early preventative therapy, 11% of children with SCD experience at least one overt ischemic stroke before the age of 20^3^, with recurrence ranging from 60-92%^4^ and mortality of roughly 5%^5^. Ischemic stroke is more common than hemorrhagic stroke in children, with hemorrhagic stroke more common in ages 20-35 years. The pathophysiology of ischemic stroke within SCD is complex^1^, and the current standard for risk assessment, transcranial doppler ultrasonography (TCD) of cerebral vessels, is difficult to perform on children under age 3. Also, some individuals with normal TCD still experience stoke^3,6,7^.

Genetic risk of stroke has been investigated in SCD and non-SCD populations. Within SCD, stroke risk is highest in *HbSS* homozygotes compared to other SCD genotypes, whereas the coinheritance of alpha-thalassemia is protective against stroke^1^. There is mounting evidence for the existence of other heritable genetic risk factors for stroke^8-15^; however, many of these are conflicting or non-replicated. A GWAS of 677 African Americans with SCD discovered two nonsynonymous variants in the genes *GOLGB1* and *ENPP1* associated with protection from stroke^13^; however, the same variant in *ENPP1* showed higher risk for stroke within a different Brazilian SCD cohort and *GOLGB1* was not replicated^15^.

Investigating genetic risk within non-SCD stroke may provide additional insight into the similarity or differences compared to SCD. Within non-SCD pediatric ischemic stroke studies, risk variants near genes *ADAMTS2, ADAMTS12*, and *ADAMTS13* haven been discovered, and both *ADAMTS2* and *ADAMTS12* have been replicated^16,17^. In separate studies, variants near *CDK18*^18^, *HABP2*^19^, were linked to early onset ischemic stroke but have not been replicated. Gaps in our knowledge of the genetic contributors to ischemic stroke risk within SCD remain. To address this, we conducted the largest genome-wide association study of stroke in individuals with sickle cell disease (N=1,333). This was conducted within the Brazil Sickle Cell Disease Cohort Study as part of the Recipient Epidemiology Donor Evaluation Study III (REDS-III)^20^.

## METHODS

Two novel approaches were implemented in this study. Variants were measured via whole genome sequencing, avoiding the need to impute on reference panels which may have little to no overlap in ancestry with the study cohort. In addition, a Cox proportional hazards model was used to estimate differences in time to stroke, allowing for the discovery of genetic variants associated with earlier stroke events.

### Study Population

The Brazil SCD Cohort Study is part of the National Institutes of Health, National Heart Lung and Blood Institute (NHLBI) REDS-III program.^21^ The Brazil National Research Ethics Commission, local ethical committees at each participating center and the Institutional Review Boards at University of California, San Francisco (UCSF) and the REDS-III data coordinating center, Research Triangle Institute, International (RTI) all reviewed and approved the study.

2,793 sickle cell disease individuals were enrolled at 6 different centers in Brazil: Hemominas Belo Horizonte, Juiz de Fora, and Montes Claros, Hemorio, Rio de Janeiro, Instituto de Tratamento do Câncer Infantil, Sao Paulo, and Hemope, Recife. Enrollment included interviews, medical abstraction, and blood collection. Written informed consent was obtained from participants ≥18 years or from guardians of younger patients, and assent was obtained for children aged 7 to 17. Medical records were abstracted for clinical history using standardized definitions^22^. This included a history of ischemic stroke defined as an acute neurological syndrome resulting from impaired cerebral blood flow without evidence of hemorrhage. Diagnostic criteria included either an MRI or CT scan showing an infarctive CNS event consistent with symptoms and signs or diagnosis based on examination and clinical history with neurologic symptoms/signs lasting >24 hours.

### DNA collection and sequencing

Whole blood was collected in EDTA, and DNA was extracted from the buffy coat via alcohol precipitation, quantified with rtPCR, and normalized to 10 ng/uL. SCD genotypes were confirmed using allele-specific pyrosequencing (Qiagen, Hilden, Germany) ^23^ and Sanger sequencing of exons 1 and 2 of *HBB* if the pyrosequencing results conflicted with medical records. Additional sequencing of *HBB* exon 3, introns, and promoter was performed for samples with unresolved genotypes at the Hemoglobinopathy Reference Laboratory at UCSF Benioff Children’s Hospital Oakland.

Whole genome sequencing was conducted within the NHLBI Trans-Omics for Precision Medicine (TOPMed) program^24^. Average coverage was 38x, and variants were called jointly across roughly 140k samples, including the N=1,333 individuals in the current study, using the GotCloud pipeline^24^ on data freeze 6a. Variants were filtered to >1% frequency, resulting in a final list of 14.3M variant autosomal sites.

### Exclusion Criteria

Individuals were excluded from this analysis for the following reasons: not having TOPMed DNA sequence data, being a duplicate sample, inability to link DNA to clinical records, outlier on PC1-3, missing age or stroke status, and not being an HbSS homozygote. Finally, we also excluded individuals with no history of stroke who were either undergoing CTT or had abnormal TCD (Suppl. Fig 1).

### Statistical Analyses

A cox proportional hazards model was used to measure SNP associations. Age at stroke was used for cases, and age at enrollment was right censored for controls without a history of ischemic stroke. Principal components (PC) were calculated from LD pruned genotypes^25^ using the entire freeze6a TOPMed cohort of roughly 140k individuals and was then was subset to just the N=1,333 participants in the study. Regressions and statistical tests were performed using the R package *survival*^*26*^ with the following model:

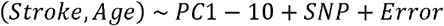

Phased genotypes were converted to integers prior to populating models as co-variates – 0 = homozygous reference; 1 = heterozygous; 2 = homozygous alternate. Genome-wide significance was defined as *P* < 5 ×10^−8^. Conditional regressions were performed on candidate 5Mb genomic regions surrounding the lead SNPs defined by lowest *P*-value. No independent loci within the candidate regions were observed. Code for this analysis is available online: https://github.com/earleyej/cox-gwas

All genotype-level analyses were performed in a HIPPA compliant environment on DNAnexus (https://www.dnanexus.com/). Annotated views of genomic candidate regions were created with LocusZoom^27^ using 1k genomes reference for linkage disequilibrium (LD). Tests on GWAS summary statistics, including gene-level association and enrichment tests were performed using the online portal Functional Mapping and Annotation of GWAS (FUMA^28^). Gene-level association was performed on the full 14M p-values using Multi-marker Analysis of GenoMic Annotation (MAGMA)^29^ using default settings. Gene set pathway analysis was performed on the Gene Ontology (GO) database (N=9,988). Colocalization analysis was performed with the R package *coloc*^*30*^ on 1Mb regions surrounding each independent genome-wide significant SNP with the lowest p-value for that region (aka ‘lead’ SNP). Additional gene set enrichment was performed on the GWAS Catalog of genes and associated conditions, as well as the Genotype-Tissue Expression (GTEx v8) databases and were performed using hypergeometric tests and corrected for multiple hypothesis testing using Benjamini-Hochberg (FDR). Candidate genes were defined as being <10kb of a lead SNP or were targets of a cis-eQTL <10kb of a lead SNP.

### Multi-variant survival analysis

Individuals were put in categories reflecting the number of lead SNPs they carried. All categories were required to have at least 10 participants, for a total of 5 categories (0, 1, 2, 3, and ≥4). These were used as categorical variables in a univariate survival analysis. *P*-values were calculated using Gehan-Wilcoxon, a non-parametric test which has more sensitivity in detecting differences between groups where the hazard ratio is higher at earlier timepoints.

## RESULTS

### Cohort characteristics

Of the 2,793 SCD individuals enrolled in the REDS-III Brazil SCD Cohort, 1,460 were excluded, with a final set of 1,333 individuals (Suppl. Fig. 1). Younger age was strongly associated with ischemic stroke (OR=0.9; CI=0.89-0.92; P<0.001; Figure 1A) but not sex (OR=0.9; CI, 0.65-1.24) or Hemocenter (OR=0.98; CI, 0.89-1.08; P=0.7). Genome-wide association was conducted on HbSS homozygotes only (N=1,333). Within this set were 178 (13.4%) individuals who experienced ischemic stroke (Table 1) with a median age at stroke of 9.5 years (Table 1, Fig 1A).

**Figure 1.**
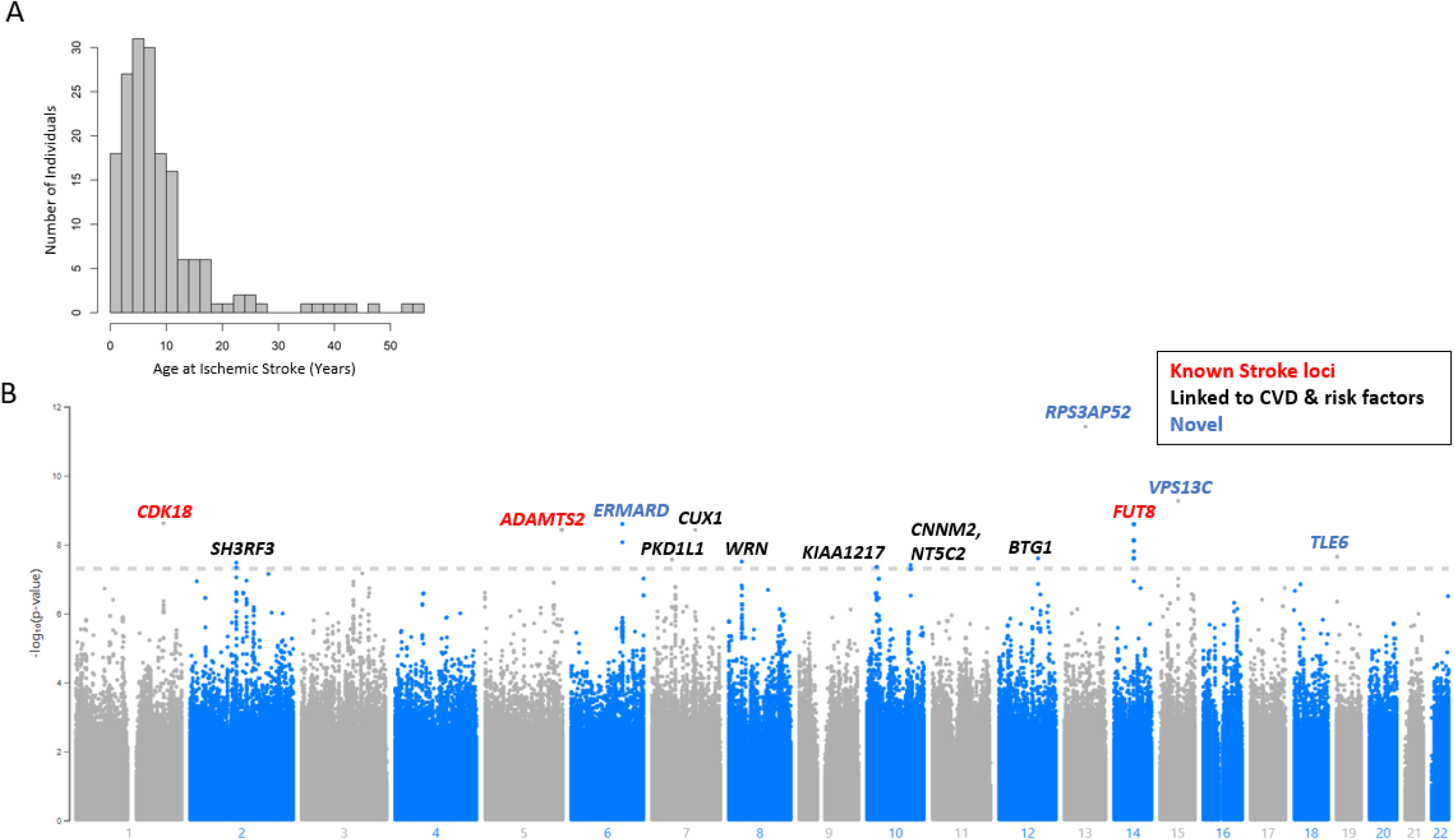
**(A)** Histogram of age at first ischemic stroke for the GWAS cohort. **(B)** Manhattan plot of results from proportional hazards GWAS P-values for ischemic stroke in HbSS SCD (N=1,333). Horizontal dotted line represents the genome-wide significance threshold of P = 5 × 10-8. The nearest gene to each lead genome-wide significant SNP is annotated with color representing confirmation of known genes linked to early onset stroke (red), genes linked to cardiovascular diseases and risk factors (black), and novel genes in blue.

**Table 1.** Demographics of HbSS Individuals within the REDS-III Brazil SCD Cohort.

|  |  |
| --- | --- |
| **Total cohort size** | **N=1,333** |
| Sex (%) |  |
| Female | 720 (54.0) |
| Male | 613 (46.0) |
| Age at enrollment, mean (±SD), y | 20.6 (±13.1) |
| Age at first stroke, mean (±SD), y | 9.5 (±9.4) |
| Stroke, no. (%) |  |
| cases | 178 (13.4) |
| controls | 1155 (86.6) |
| Hemocenter, no. (%) |  |
| Hemominas BH | 313 (23.5) |
| Hemominas JFO | 149 (11.2) |
| Hemominas MOC | 205 (15.4) |
| Hemope | 285 (21.4) |
| Hemorio | 332 (24.9) |
| ITACI SP | 49 (0.04) |

### Genetic architecture of ischemic stroke in Brazilian Sickle Cell

We observed 28 genome-wide significant variants (Suppl. Table 1) across 14 independent regions associated with stroke occurring at younger ages (Figure 1B, Table 2, Suppl. Fig 2). No genomic inflation in *P*-values was observed (Supplemental Figure 3). Most variants sites were intergenic; however, three regions were intronic to genes *CUX1, KIAA1217*, and *CNNM2*. Also Included in this list of 14 lead SNPs were three regions previously linked to stroke – *ADAMTS2, CDK18, FUT8*^17-19^.

**Table 2.**
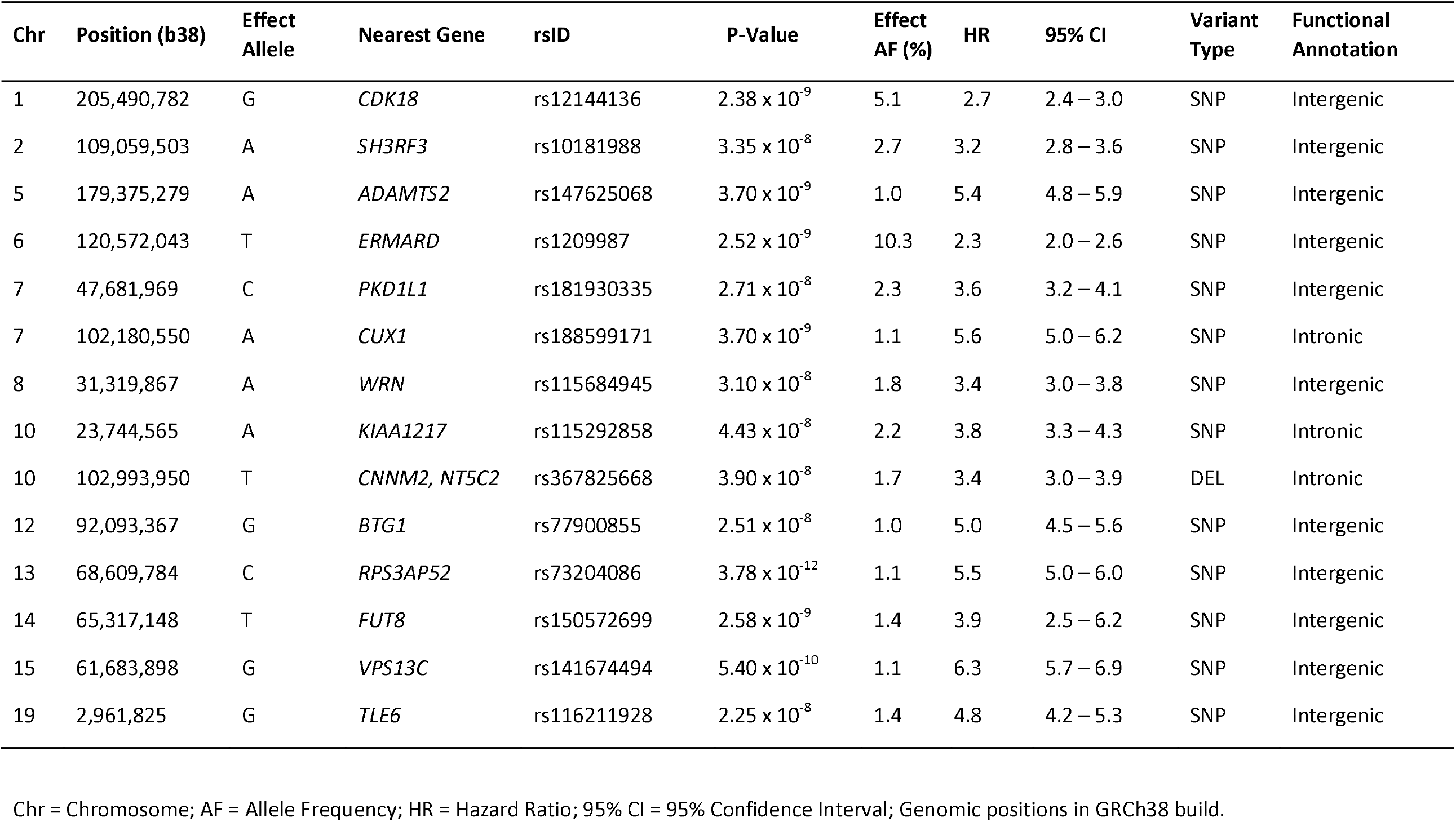
Cox regression results for lead genome-wide significant SNPs.

Out of the 14 genome-wide lead variants, 5 were only observed in heterozygous state whereas 9 were observed in both heterozygous and homozygous variant state. Individuals possessing at least one of these 14 variants exhibited increased risk for ischemic stroke at earlier ages (P < 0.001 for all 14 variants, Gehan-Wilcoxon) (Suppl Figure 4).

### Multiple variant survival analysis

Individuals with zero lead SNPs had a stroke rate of 4% (n=716), whereas those with at least one of the 14 lead variants had an increased rate at 14% (n=430), and this rate increased even further for individuals with two (36%; n=125), three (70%; n=50), or four or more variants (83%; n=12), and this trend was significant (*P* < 0.001, Gehan-Wilcoxon; Figure 2).

**Figure 2.**
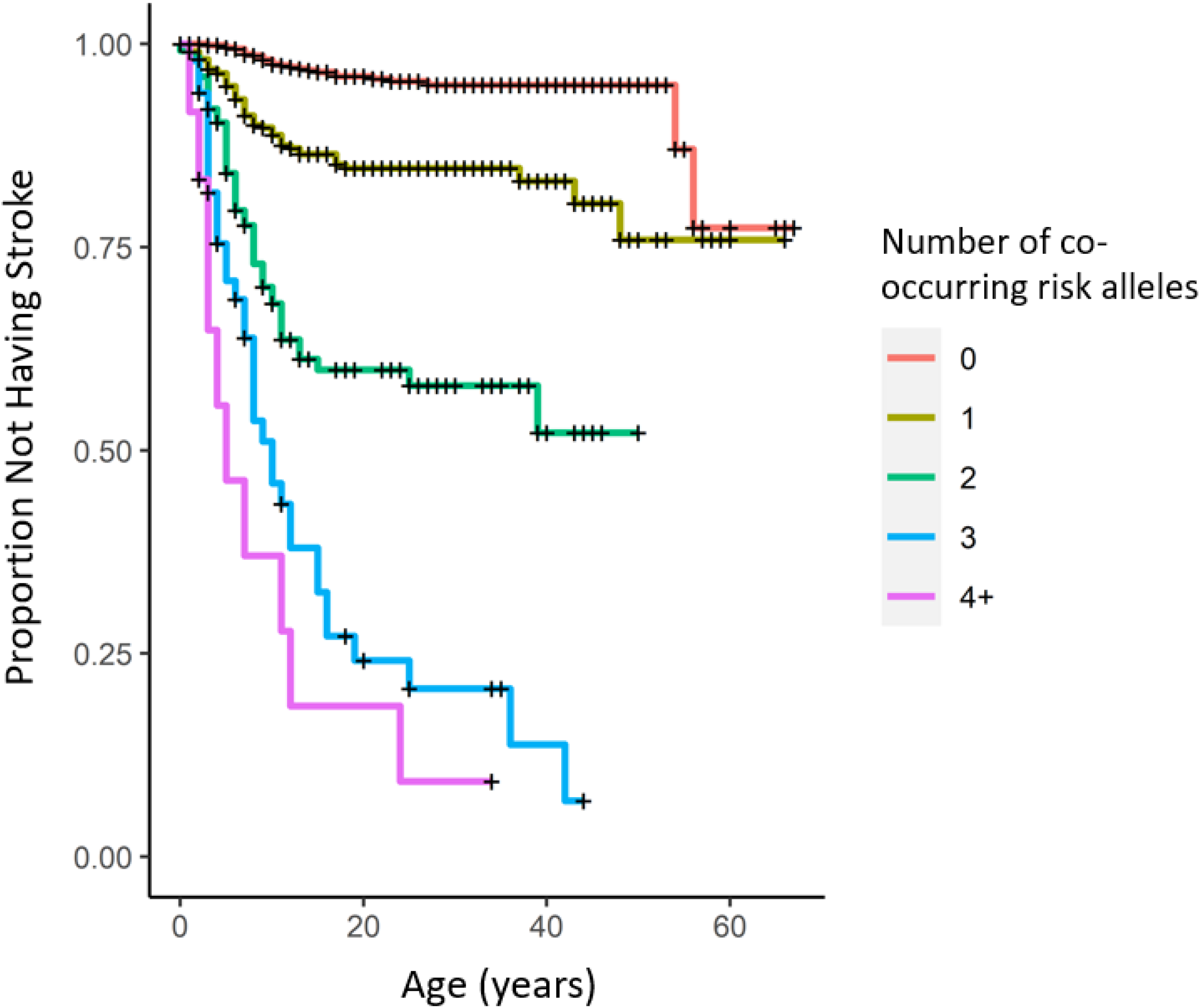
Kaplan-Meier plot of individuals harboring multiple co-occurring genome-wide significant risk alleles for early stroke. Curves represent subsets of the study cohort who possessed zero (red, N=716), one (olive, N=430), two (green, N=125), three (blue, N=50), or four or more (purple, N=12) stroke risk alleles in any combination from the 14 lead SNPS. (+) represents right-censored data.

### Cis-co-localization analysis of ischemic stroke using GTEx

SNPs within the 14 candidate regions were then searched against the GTEx database of cis-eQTL variant associations across different tissue types. The lead SNP on chromosome 1, rs12144136, is a *cis*-eQTL for the gene *CDK18* within the Skeletal Muscle tissue category (Suppl. Table 2) which appears to upregulate this gene. No other SNPs at genome-wide significance showed evidence of *cis*-eQTL activity.

We assessed co-localization of cis-eQTL associations with 24 different tissue types relevant to stroke. Three of the 14 candidate regions showed strong evidence (PP > 0.9) of colocalization with a *cis*-eQTL (Suppl. Table 2). On chromosome 12, two *cis*-eQTLs were found for genes *LUM* in the frontal cortex (rs77217583) and *NUDT4* (rs117881990) in the hypothalamus. On chromosome 14, one *cis*-eQTL was found for *RAB15* in the substantia nigra; and on chromosome 15, one *cis*-eQTL was found for *VPS13C* in cervical spinal cord. Three other moderately strong signals (P > 0.5) were detected on chromosome 8 – one *cis*-eQTL for *WRN* expression in heart left ventricle and two *cis*-eQTLs for *PURG* in the substantia nigra.

### Gene-level Association

We performed a gene-level association on 19,021 protein coding genes using the 14M site summary statistics (Supplemental Figure 5; Supplementary Table 3). No genes exhibited genome-wide significant enrichment defined at the Bonferroni-corrected alpha of 2.6×10^−6^; however, three genes exhibited some evidence of association (*P*<10^−4^), including *GPR15* (P=3.9×10-^5^), *MMP26* (P=4.6×10^−5^), and *GOLT1B* (P=7.5×10-^5^).

Gene-set enrichment analysis was performed to predict impact on biological functions using the Gene Ontology (GO) database (Supplemental Table 4). While no gene set achieved the Bonferroni-corrected p-value of 5×10^−6^, top results at the *P*<0.001 threshold included the platelet dense tubular network (*P* = 3.63×10^−5^), the hemoglobin complex (*P* = 0.0007), gas transport (*P* = 0.0006), and regulation of systemic arterial blood pressure by hormone (*P* = 0.0008).

### Candidate-gene Enrichment Analysis

To assess potential tissue-specific expression, enrichment tests were performed on the GTEx catalog using 18 candidate genes selected using two criteria: located <10kb of a lead SNP; or target of an eQTL located <10kb of a lead SNP (Suppl. Table 5). Significant enrichment was observed in multiple tissue classes, including upregulated genes in the coronary (8 genes, *P* = 0.0005, FDR), tibial (9, *P* = 0.03, FDR) and aorta arteries (7, *P* = 0.03, FDR), as well as downregulated genes in the spleen (7, *P* = 0.005), pancreas (13 genes, *P* = 0.02), esophagus mucosa (8, *P* = 0.03), hypothalamus (10, *P* = 0.03), and substantia nigra (11, *P* = 0.03) (Figure 3A, Suppl. Table 6).

**Figure 3.**
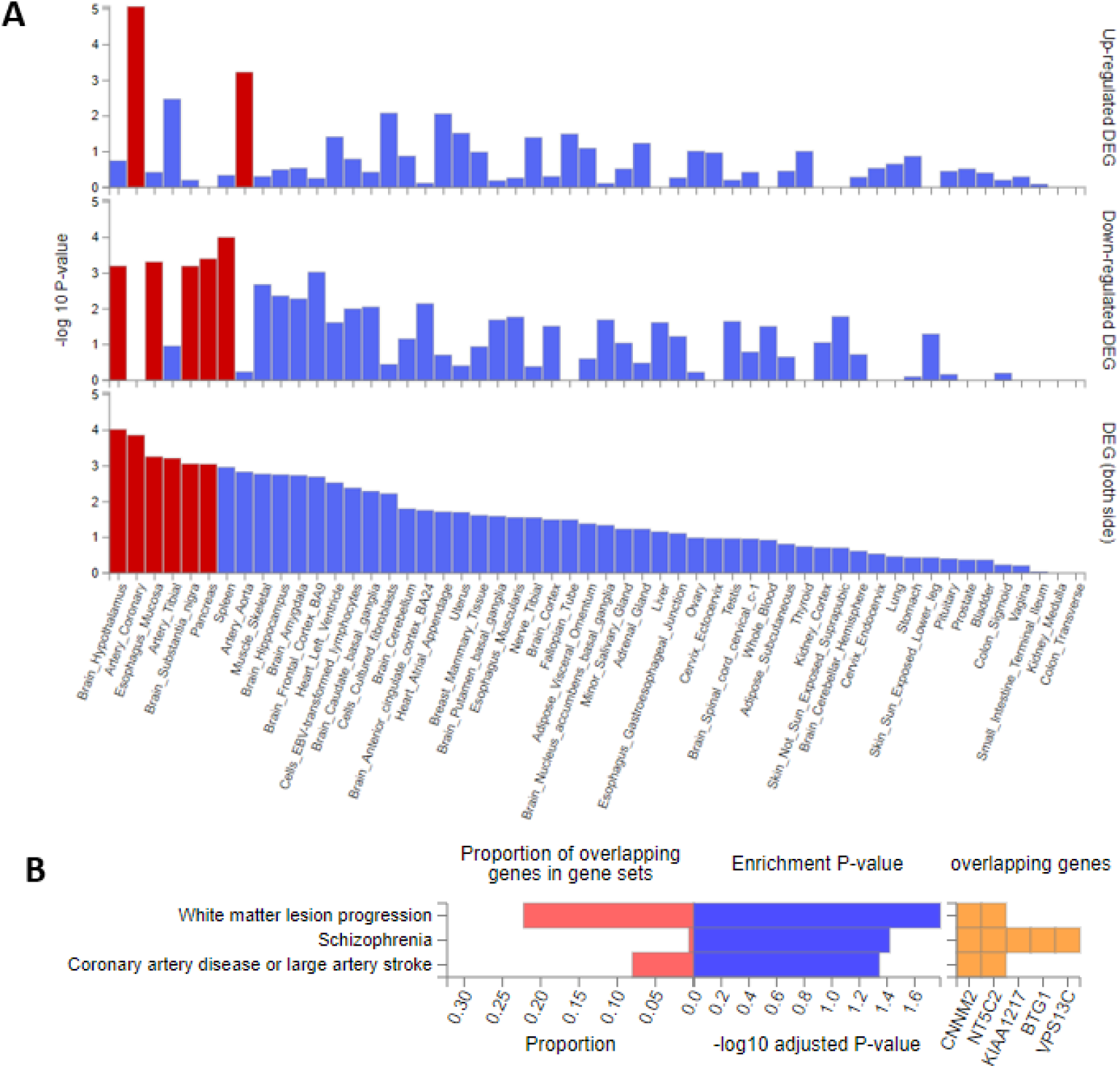
Gene set enrichment test results for 18 candidate genes. (A) Barplots of transformed P-values (FDR) from hypergeometric enrichment test of 18 genes located within 10kb of a lead SNP (15 genes) or were predicted targets of a cis-eQTL in the same region (3 genes). Red bars show tissues observed at P < 0.05. Left panel, general tissue classes; right panel, specific tissue types. (B) Horizontal barplots of significant enrichment results against the GWAS Catalog. Red, proportion of genes; blue, transformed p-values (FDR); yellow, genes.

Gene enrichment test using the GWAS Catalog uncovered significant associations to disease categories of white matter lesion progression (*CNNM2, NT5C2, P* = 0.01) and coronary artery disease/large artery stroke (*CNNM2, NT5C2, P* = 0.04)(Figure 3B). A second enrichment test was performed on an expanded list of 35 genes (<100kb of a lead SNP), and this set exhibited significant overlap with genes linked to hypertension (5 genes, *P* = 7.8×10^−7^), white matter lesion progression (3 genes, *P* = 4.67×10^−7^), mean arterial pressure (8, *P* = 7.81×10^−11^), microalbuminuria (4, *P* = 1.86×10^−7^), coronary artery disease (7, *P* = 8.93×10^−6^), and coronary artery disease/large artery stroke (2, *P* = 0.0007) (Suppl. Fig 6).

### Comparison to previous ischemic stroke studies in SCD and non-SCD

Previous candidate SNP studies of stroke in SCD have evaluated 12 SNPs, as well as one additional SNP from a GWAS^8-15^. None of these candidates SNPs reached genome-wide significance in the current study (Suppl. Table 7). Expanding the search to surrounding 1Mb region centered on the 13 candidate SNPs uncovered the lead SNP rs147625068 (*P* = 3.7×10^−9^) near gene *LTC4S* on chromosome 9.

A similar search was performed for non-SCD early onset ischemic stroke candidate genes: *ADAMTS2* (rs469568), *ADAMTS12* (rs1364044), *CDK18* (rs77571454), *HABP2 (*rs11196288) ^16-19^. In the current study, the variant rs77571454 near gene *CDK18* was not observed, and none of the other three variants reached genome-wide significance (Suppl Table 8). Searching within the 1Mb region centered on these four candidate SNPs uncovered the two lead SNPs rs12144136 (*P* = 2.38×10^−9^), near *CDK18*, and rs147625068 (*P* = 3.7×10^−9^), near *ADAMTS2*, and one additional variant near *ADAMTS12* (rs534089428, *P* = 2.43×10^−5^).

## DISCUSSSION

We have discovered 14 genetic regions associated with ischemic stroke occurring at earlier ages in SCD individuals with homozygous HbSS genotype, encompassing 18 candidate genes. This study included 1,333 HbSS individuals, the largest genome-wide evaluation of ischemic stroke within SCD to date. We implemented a cox proportional hazards model to estimate risk at earlier timepoints. Results included gene candidates previously linked with non-SCD early-onset stroke – *ADAMTS2, CDK18*; genes linked to cardiovascular disease risk factors – *BTG1, CNMM2, CUX1, EMARD, FUT8, KIAA1217, LUM, NT5C2, NUDT4, PKD1L1, WRN*; and eight novel genes – *SH3RF3, RAB15, RPS3AP52, VPS13C*, and *TLE6*. To our knowledge this is the first independent replication of *CDK18* and *FUT8*.

Previous candidate gene studies of stroke in SCD replicated five alleles within genes *TGFBR3, TEK, ANXA2, IL4R*, and *ADCY9*^10-12^; however, the single published GWAS of ischemic stroke in SCD did not replicate these and instead proposed two novel genes – *GOLGB1* and *ENPP1*^13^. We did not observe replication of the candidate genes nor the GWAS variants. The reasons for this are unclear, although we should note the previous GWAS enrolled African Americans whereas the current work enrolled Brazilians, each having distinct histories of admixture.

Results from this work replicate stroke risk genes for non-SCD early onset stroke: *ADAMTS2* and *CDK18*^16-18^. Members of the *ADAMTS* gene family, in particular *ADAMTS2* and *ADAMTS12*, have been linked to pediatric stroke in two separate U.S. populations^16,17^. *ADAMTS2* also appears to have a role in cerebral aneurysm^31^ and cardiac muscle hypertrophy^32^ possibly through regulation of the extracellular matrix^33^. We did observe one variant, rs534089428, within 500kb of *ADAMTS12* at *P* = 2×10^−5^. *CDK18* has not been replicated until now.

High reticulocyte count is a reliable risk factor for ischemic stroke in sickle cell individuals^1^. Candidate genes from the current study, *CUX1, CNNM2*, and *NUDT4*, have been previously associated with higher reticulocyte counts^34 35^. *CNNM2* has also been linked to numerous stroke related traits – hypertension, coronary artery disease, and mean corpuscular hemoglobin ^36-43^. *CNNM2* gene is expressed throughout the body but is particularly high in the brain, kidney, and endocrine tissue^44^ and appears to play a role in renal magnesium uptake^45,46^

Pathway analysis also uncovered enrichment of variants in genes involved with the platelet dense tubular network. Platelets have been shown to play a role in SCD pathophysiology^47-49^, and higher platelet aggregation and hypercoagulability has been observed in blood from non-SCD ischemic stroke patients^50^. Results from the current work suggest a systemic dysregulation of platelet activation-sequestration within SCD children suffering from strokes, but further studies will be required to confirm this.

In conclusion, we have identified risk alleles associated with earlier ischemic stroke. Findings from this work have confirmed previously identified non-SCD early onset ischemic stroke genes – *ADAMTS2* and *CDK18*– and discovered 16 novel genes. Future studies will be needed to disentangle the potentially complex interaction among these alleles.

## Data Availability

Molecular data and associated clinical and phenotype data are available within dbGaP https://www.ncbi.nlm.nih.gov/projects/gap/cgi-bin/study.cgi?study_id=phs001599.v1.p1

https://www.ncbi.nlm.nih.gov/projects/gap/cgi-bin/study.cgi?study_id=phs001599.v1.p1

## Acknowledgements

Special thanks to patients and research staff at each hemocenter in Brazil participating in this study. We recognize the following people for their commitment and contribution to this project: Alfredo Mendrone Jr., Cesar de Almeida Neto, Roberta Calcucci, Erivanda Bezerra, Carolina Miranda, Franciane Mendes de Oliveira, Valquıria Reis, Nayara Durte, Barbara Malta, Jose Wilson Sales, Maria Aparecida Souza, Rodrigo Ferreira, Maria do Carmo Valgueir, Regina Gomes, Airly Goes Maciel, Rebeca Talamatu Dantas, Flavia Herculano, Ana Claudia Pereira, Ana Carla Alvarenga, Adriana Grilo, Fabiana Canedo, Pedro Losco Takecian, Mina Cintho Ozahata, Rodrigo Muller de Carvalho, Christopher McClure, Simone A. Glynn.

Molecular data for the Trans-Omics in Precision Medicine (TOPMed) program was supported by the National Heart, Lung and Blood Institute (NHLBI). Genome Sequencing for “NHLBI TOPMed: REDS-III_Brazil” (phs001468.v3.p1) was performed at Baylor (HHSN268201500015C, HHSN268201600033I). Core support including centralized genomic read mapping and genotype calling, along with variant quality metrics and filtering were provided by the TOPMed Informatics Research Center (3R01HL-117626-02S1; contract HHSN268201800002I). Core support including phenotype harmonization, data management, sample-identity QC, and general program coordination were provided by the TOPMed Data Coordinating Center (R01HL-120393; U01HL-120393; contract HHSN268201800001I). We gratefully acknowledge the studies and participants who provided biological samples and data for TOPMed.

## REFERENCES

1. Belisario AR, Silva CM, Velloso-Rodrigues C, Viana MB. Genetic, laboratory and clinical risk factors in the development of overt ischemic stroke in children with sickle cell disease. Hematol Transfus Cell Ther. 2018;40(2):166–181.

2. Earley CJ, Kittner SJ, Feeser BR, et al. Stroke in children and sickle-cell disease: Baltimore-Washington Cooperative Young Stroke Study. Neurology. 1998;51(1):169–176.

3. Ohene-Frempong K, Weiner SJ, Sleeper LA, et al. Cerebrovascular accidents in sickle cell disease: rates and risk factors. Blood. 1998;91(1):288–294.

4. Kirkham FJ, Lagunju IA. Epidemiology of Stroke in Sickle Cell Disease. J Clin Med. 2021;10(18).

5. Strouse JJ, Jordan LC, Lanzkron S, Casella JF. The excess burden of stroke in hospitalized adults with sickle cell disease. Am J Hematol. 2009;84(9):548–552.

6. Adams RJ, McKie VC, Carl EM, et al. Long-term stroke risk in children with sickle cell disease screened with transcranial Doppler. Ann Neurol. 1997;42(5):699–704.

7. Adams RJ, Brambilla DJ, Granger S, et al. Stroke and conversion to high risk in children screened with transcranial Doppler ultrasound during the STOP study. Blood. 2004;103(10):3689–3694.

8. Hoppe C, Klitz W, Cheng S, et al. Gene interactions and stroke risk in children with sickle cell anemia. Blood. 2004;103(6):2391–2396.

9. Taylor JGt, Tang DC, Savage SA, et al. Variants in the VCAM1 gene and risk for symptomatic stroke in sickle cell disease. Blood. 2002;100(13):4303–4309.

10. Sebastiani P, Ramoni MF, Nolan V, Baldwin CT, Steinberg MH. Genetic dissection and prognostic modeling of overt stroke in sickle cell anemia. Nat Genet. 2005;37(4):435–440.

11. Flanagan JM, Frohlich DM, Howard TA, et al. Genetic predictors for stroke in children with sickle cell anemia. Blood. 2011;117(24):6681–6684.

12. Belisario AR, Sales RR, Toledo NE, et al. Reticulocyte count is the most important predictor of acute cerebral ischemia and high-risk transcranial Doppler in a newborn cohort of 395 children with sickle cell anemia. Ann Hematol. 2016;95(11):1869–1880.

13. Flanagan JM, Sheehan V, Linder H, et al. Genetic mapping and exome sequencing identify 2 mutations associated with stroke protection in pediatric patients with sickle cell anemia. Blood. 2013;121(16):3237–3245.

14. Hoppe C, Klitz W, D’Harlingue K, et al. Confirmation of an association between the TNF(−308) promoter polymorphism and stroke risk in children with sickle cell anemia. Stroke. 2007;38(8):2241–2246.

15. Belisario AR, Nogueira FL, Rodrigues RS, et al. Association of alpha-thalassemia, TNF-alpha (−308GA) and VCAM-1 (c.1238GC) gene polymorphisms with cerebrovascular disease in a newborn cohort of 411 children with sickle cell anemia. Blood Cells Mol Dis. 2015;54(1):44–50.

16. Witten A, Ruhle F, de Witt M, et al. ADAMTS12, a new candidate gene for pediatric stroke. PLoS One. 2020;15(8):e0237928.

17. Arning A, Hiersche M, Witten A, et al. A genome-wide association study identifies a gene network of ADAMTS genes in the predisposition to pediatric stroke. Blood. 2012;120(26):5231–5236.

18. Yamada Y, Kato K, Oguri M, et al. Identification of nine genes as novel susceptibility loci for early-onset ischemic stroke, intracerebral hemorrhage, or subarachnoid hemorrhage. Biomed Rep. 2018;9(1):8–20.

19. Cheng YC, Stanne TM, Giese AK, et al. Genome-Wide Association Analysis of Young-Onset Stroke Identifies a Locus on Chromosome 10q25 Near HABP2. Stroke. 2016;47(2):307–316.

20. Carneiro-Proietti ABF, Kelly S, Miranda Teixeira C, et al. Clinical and genetic ancestry profile of a large multi-centre sickle cell disease cohort in Brazil. Br J Haematol. 2018;182(6):895–908.

21. Kleinman S, Busch MP, Murphy EL, et al. The National Heart, Lung, and Blood Institute Recipient Epidemiology and Donor Evaluation Study (REDS-III): a research program striving to improve blood donor and transfusion recipient outcomes. Transfusion. 2014;54(3 Pt 2):942–955.

22. Ballas SK, Lieff S, Benjamin LJ, et al. Definitions of the phenotypic manifestations of sickle cell disease. Am J Hematol. 2010;85(1):6–13.

23. de Martino CC, Alencar CS, Loureiro P, et al. Use of an automated pyrosequencing technique for confirmation of sickle cell disease. PLoS One. 2019;14(12):e0216020.

24. Taliun D, Harris DN, Kessler MD, et al. Sequencing of 53,831 diverse genomes from the NHLBI TOPMed Program. Nature. 2021;590(7845):290–299.

25. Purcell S, Neale B, Todd-Brown K, et al. PLINK: a tool set for whole-genome association and population-based linkage analyses. Am J Hum Genet. 2007;81(3):559–575.

26. A Package for Suvival Analysis in R. [computer program]. 2020.

27. Pruim RJ, Welch RP, Sanna S, et al. LocusZoom: regional visualization of genome-wide association scan results. Bioinformatics. 2010;26(18):2336–2337.

28. Watanabe K, Taskesen E, van Bochoven A, Posthuma D. Functional mapping and annotation of genetic associations with FUMA. Nat Commun. 2017;8(1):1826.

29. de Leeuw CA, Mooij JM, Heskes T, Posthuma D. MAGMA: generalized gene-set analysis of GWAS data. PLoS Comput Biol. 2015;11(4):e1004219.

30. Giambartolomei C, Vukcevic D, Schadt EE, et al. Bayesian test for colocalisation between pairs of genetic association studies using summary statistics. PLoS Genet. 2014;10(5):e1004383.

31. Arning A, Jeibmann A, Kohnemann S, et al. ADAMTS genes and the risk of cerebral aneurysm. J Neurosurg. 2016;125(2):269–274.

32. Wang X, Chen W, Zhang J, et al. Critical Role of ADAMTS2 (A Disintegrin and Metalloproteinase With Thrombospondin Motifs 2) in Cardiac Hypertrophy Induced by Pressure Overload. Hypertension. 2017;69(6):1060–1069.

33. Mead TJ, Apte SS. ADAMTS proteins in human disorders. Matrix Biol. 2018;71-72:225–239.

34. Astle WJ, Elding H, Jiang T, et al. The Allelic Landscape of Human Blood Cell Trait Variation and Links to Common Complex Disease. Cell. 2016;167(5):1415–1429 e1419.

35. Vuckovic D, Bao EL, Akbari P, et al. The Polygenic and Monogenic Basis of Blood Traits and Diseases. Cell. 2020;182(5):1214–1231 e1211.

36. Wain LV, Vaez A, Jansen R, et al. Novel Blood Pressure Locus and Gene Discovery Using Genome-Wide Association Study and Expression Data Sets From Blood and the Kidney. Hypertension. 2017.

37. Takeuchi F, Akiyama M, Matoba N, et al. Interethnic analyses of blood pressure loci in populations of East Asian and European descent. Nat Commun. 2018;9(1):5052.

38. German CA, Sinsheimer JS, Klimentidis YC, Zhou H, Zhou JJ. Ordered multinomial regression for genetic association analysis of ordinal phenotypes at Biobank scale. Genet Epidemiol. 2020;44(3):248–260.

39. Jeong H, Jin HS, Kim SS, Shin D. Identifying Interactions between Dietary Sodium, Potassium, Sodium-Potassium Ratios, and FGF5 rs16998073 Variants and Their Associated Risk for Hypertension in Korean Adults. Nutrients. 2020;12(7).

40. Wu TH, Fann JC, Chen SL, et al. Gradient Relationship between Increased Mean Corpuscular Volume and Mortality Associated with Cerebral Ischemic Stroke and Ischemic Heart Disease: A Longitudinal Study on 66,294 Taiwanese. Sci Rep. 2018;8(1):16517.

41. van der Harst P, Verweij N. Identification of 64 Novel Genetic Loci Provides an Expanded View on the Genetic Architecture of Coronary Artery Disease. Circ Res. 2018;122(3):433–443.

42. Dichgans M, Malik R, Konig IR, et al. Shared genetic susceptibility to ischemic stroke and coronary artery disease: a genome-wide analysis of common variants. Stroke. 2014;45(1):24–36.

43. Koyama S, Ito K, Terao C, et al. Population-specific and trans-ancestry genome-wide analyses identify distinct and shared genetic risk loci for coronary artery disease. Nat Genet. 2020;52(11):1169–1177.

44. Uhlen M, Fagerberg L, Hallstrom BM, et al. Proteomics. Tissue-based map of the human proteome. Science. 2015;347(6220):1260419.

45. Arjona FJ, de Baaij JH, Schlingmann KP, et al. CNNM2 mutations cause impaired brain development and seizures in patients with hypomagnesemia. PLoS Genet. 2014;10(4):e1004267.

46. Hartmann K, Seweryn M, Handelman SK, Rempala GA, Sadee W. Non-linear interactions between candidate genes of myocardial infarction revealed in mRNA expression profiles. BMC Genomics. 2016;17(1):738.

47. Inwald DP, Kirkham FJ, Peters MJ, et al. Platelet and leucocyte activation in childhood sickle cell disease: association with nocturnal hypoxaemia. Br J Haematol. 2000;111(2):474–481.

48. Westwick J, Watson-Williams EJ, Krishnamurthi S, et al. Platelet activation during steady state sickle cell disease. J Med. 1983;14(1):17–36.

49. Wun T, Paglieroni T, Rangaswami A, et al. Platelet activation in patients with sickle cell disease. Br J Haematol. 1998;100(4):741–749.

50. Tutwiler V, Peshkova AD, Andrianova IA, Khasanova DR, Weisel JW, Litvinov RI. Contraction of Blood Clots Is Impaired in Acute Ischemic Stroke. Arterioscler Thromb Vasc Biol. 2017;37(2):271–279.

