## Supplemental Material for "Genome-wide association study of ischemic stroke risk in Sickle Cell confirms *ADAMTS2, CDK18*, uncovers 12 novel loci"

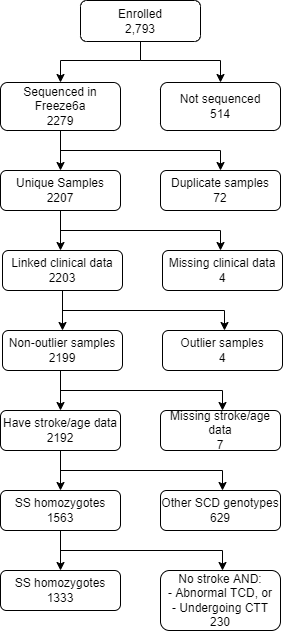

**Supplemental Figure 1.** Consort diagram of participants within the current study. The GWAS was performed on HbSS homozygotes (N=1,333). SCD: sickle cell disease; TCD: transcranial doppler; CTT: chronic transfusion therapy; freeze6a: the genotype data version.

**Supplemental Table 1.** All genome-wide significant SNPs at *P* < 5x10^-8^

| Chr | Pos | Ref | Alt | rsID | HR | 95% CI | P | MAF |
| --- | --- | --- | --- | --- | --- | --- | --- | --- |
| chr1 | 205490782 | T | G | rs12144136 | 2.7 | 2.4 – 3.0 | 2.38E-09 | 0.051 |
| chr2 | 109016367 | G | A | rs144267740 | 4.2 | 3.7 – 4.7 | 4.63E-08 | 0.017 |
| chr2 | 109059503 | G | A | rs10181988 | 3.2 | 2.8 – 3.6 | 3.35E-08 | 0.027 |
| chr5 | 179375279 | G | A | rs147625068 | 5.4 | 4.8 – 5.9 | 3.70E-09 | 0.010 |
| chr5 | 179377185 | G | A | rs113596294 | 5.4 | 4.8 – 5.9 | 3.70E-09 | 0.010 |
| chr5 | 179378831 | A | G | rs111696014 | 5.4 | 4.8 – 5.9 | 3.70E-09 | 0.010 |
| chr6 | 120565197 | A | T | rs2207754 | 2.3 | 2.0 – 2.6 | 8.52E-09 | 0.101 |
| chr6 | 120572043 | T | C | rs1209987 | 2.3 | 2.0 – 2.6 | 2.52E-09 | 0.103 |
| chr7 | 47681969 | T | C | rs181930335 | 3.6 | 3.2 – 4.1 | 2.71E-08 | 0.023 |
| chr7 | 102180550 | G | A | rs188599171 | 5.6 | 5.0 – 6.2 | 3.70E-09 | 0.011 |
| chr8 | 31319867 | C | A | rs115684945 | 3.4 | 3.0 – 3.8 | 3.10E-08 | 0.018 |
| chr10 | 23744565 | G | A | rs115292858 | 3.8 | 3.3 – 4.3 | 4.43E-08 | 0.022 |
| chr10 | 102993950 | TGA | T | rs367825668 | 3.4 | 3.0 – 3.9 | 3.90E-08 | 0.017 |
| chr12 | 92093367 | A | G | rs77900855 | 5.0 | 4.5 – 5.6 | 2.51E-08 | 0.010 |
| chr13 | 68609784 | T | C | rs73204086 | 5.5 | 5.0 – 6.0 | 3.78E-12 | 0.011 |
| chr14 | 65293428 | A | G | rs144177699 | 3.8 | 3.4 – 4.3 | 7.56E-09 | 0.014 |
| chr14 | 65293545 | C | T | rs141155259 | 3.8 | 3.4 – 4.3 | 7.56E-09 | 0.014 |
| chr14 | 65294188 | T | C | rs140265978 | 3.8 | 3.4 – 4.3 | 7.56E-09 | 0.014 |
| chr14 | 65295693 | C | T | rs139331229 | 3.6 | 3.2 – 4.1 | 1.57E-08 | 0.016 |
| chr14 | 65298256 | A | G | rs115143940 | 3.8 | 3.4 – 4.3 | 7.56E-09 | 0.014 |
| chr14 | 65302668 | G | A | rs116767851 | 3.8 | 3.3 – 4.3 | 2.50E-08 | 0.014 |
| chr14 | 65305710 | A | G | rs185065699 | 3.8 | 3.3 – 4.3 | 2.50E-08 | 0.014 |
| chr14 | 65317148 | G | T | rs150572699 | 3.9 | 3.5 – 4.4 | 2.58E-09 | 0.014 |
| chr14 | 65317441 | C | T | rs141663600 | 3.9 | 3.5 – 4.4 | 2.58E-09 | 0.014 |
| chr14 | 65323254 | G | A | rs147928562 | 3.8 | 3.3 – 4.3 | 2.50E-08 | 0.014 |
| chr14 | 65323725 | T | C | rs146742007 | 3.9 | 3.5 – 4.4 | 2.58E-09 | 0.014 |
| chr15 | 61683898 | A | G | rs141674494 | 6.3 | 5.7 – 6.9 | 5.40E-10 | 0.011 |
| chr19 | 2961825 | T | G | rs116211928 | 4.8 | 4.2 – 5.3 | 2.25E-08 | 0.014 |

Chr = chromosome; Pos = position (hg38); Ref = reference allele; Alt = Alternate allele; MAF = minor allele frequency; HR = hazards ratio; 95% CI = 95% confidence interval

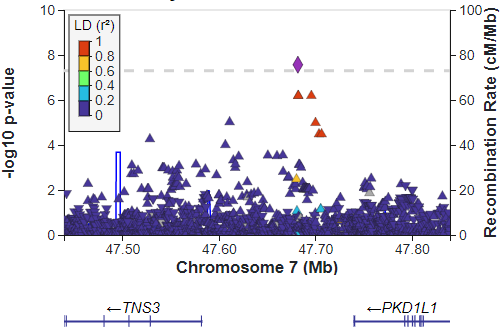

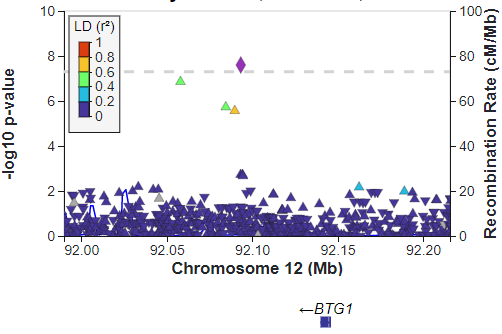

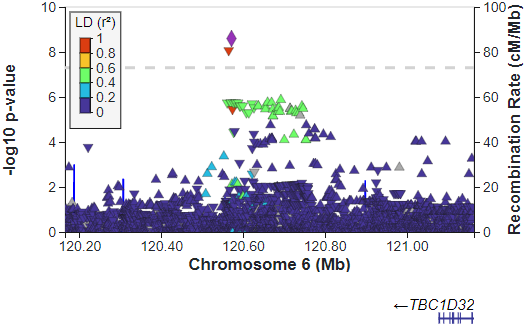

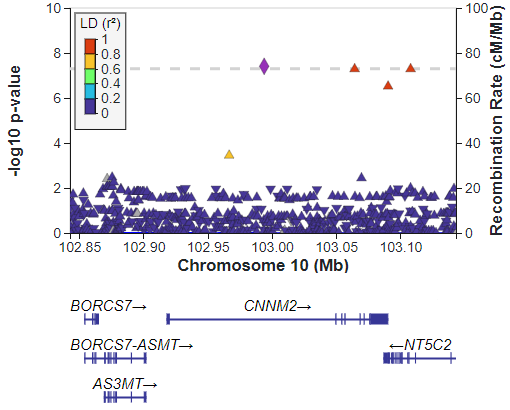

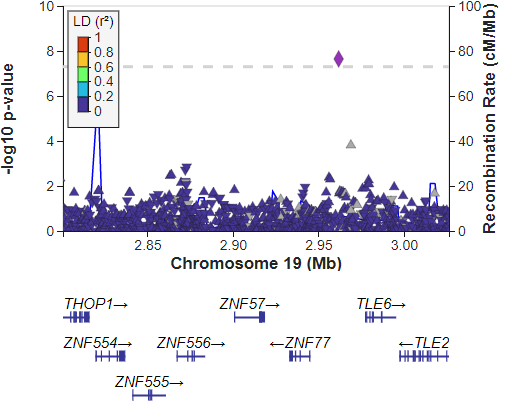

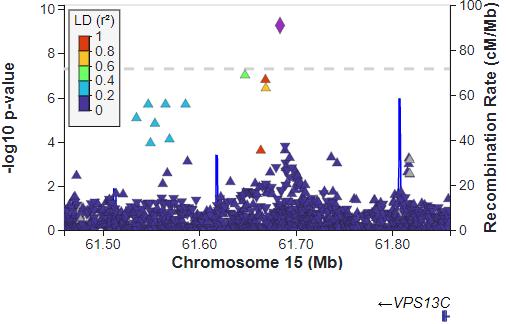

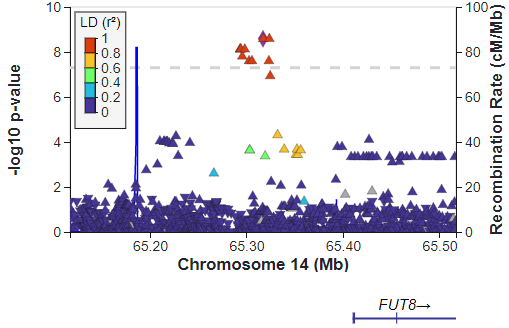

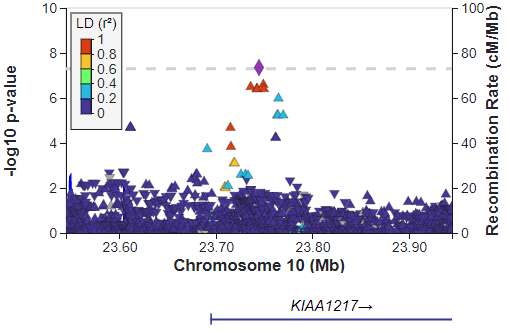

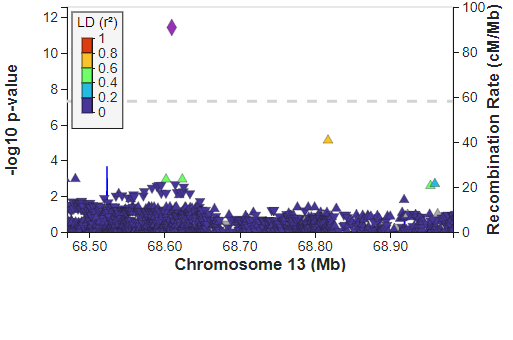

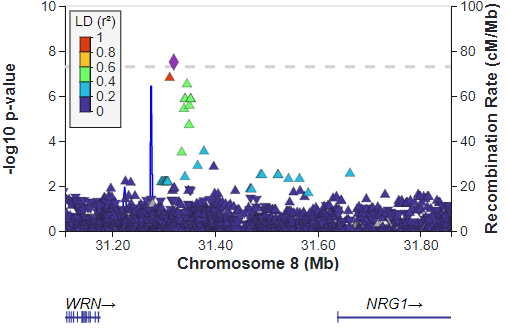

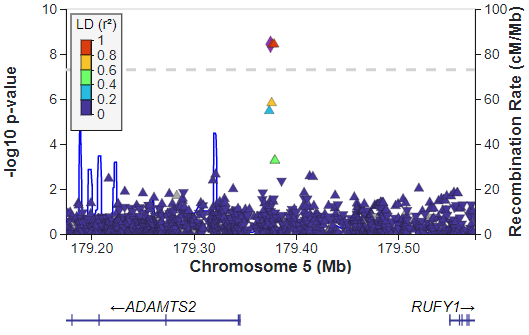

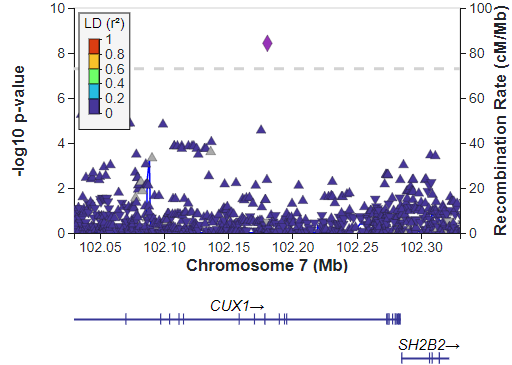

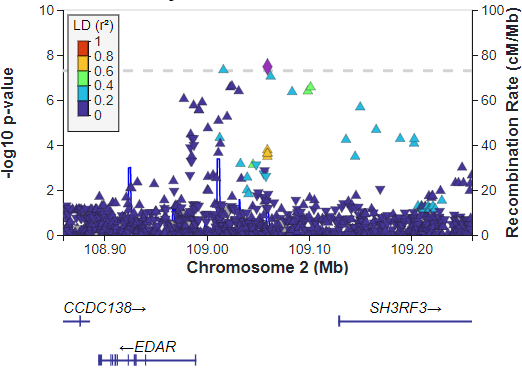

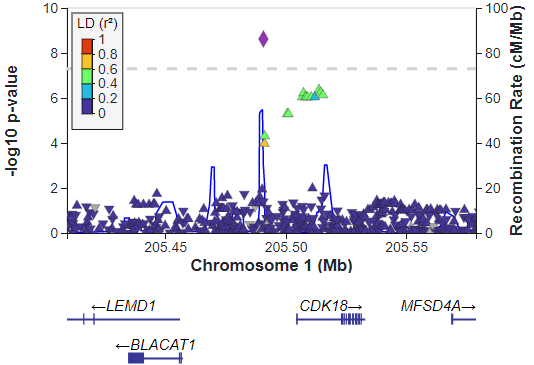

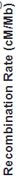

**Supplemental Figure 2.** LocusZoom plots of candidate regions reaching genome-wide significance (gray dotted line, *P* < 5 x 10^-8^). Purple diamonds represent lead SNPs. Triangles show -log10 transformed p-values across all observed positions, with the direction of the triangle representing direction of estimated allelic effect with color representing linkage (*r^2^*) to the lead SNP. Blue lines within panels represent recombination rates (cM/Mb). 1000 genomes phase3 (“ALL”) was used as reference to calculate LD and recombination rates. Gene models are shown below each panel to give genomic context

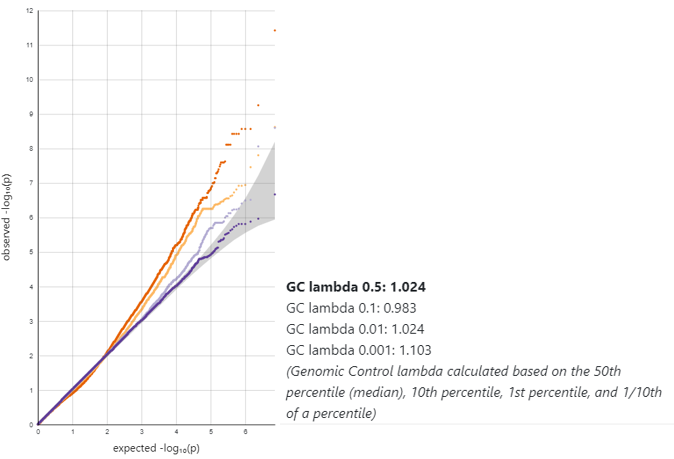

**Supplemental Figure 3.** QQ plot of observed versus expected p-values and lambda values at decreasing allele frequencies.

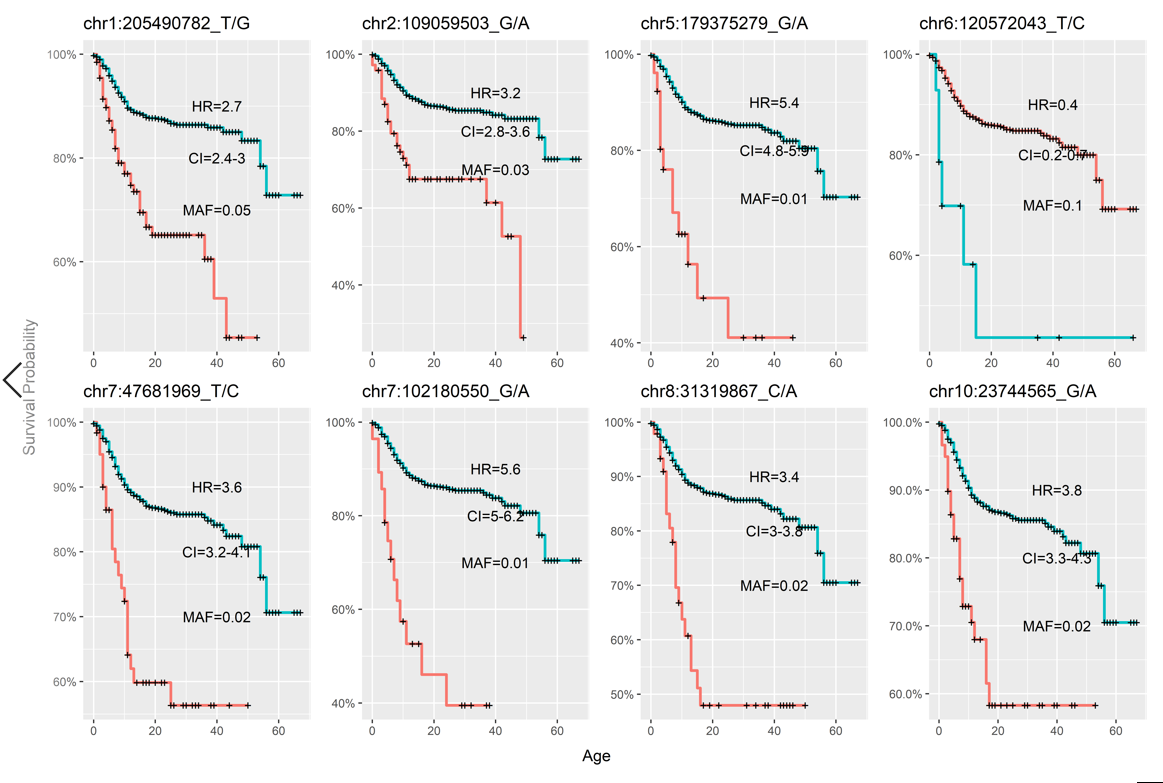

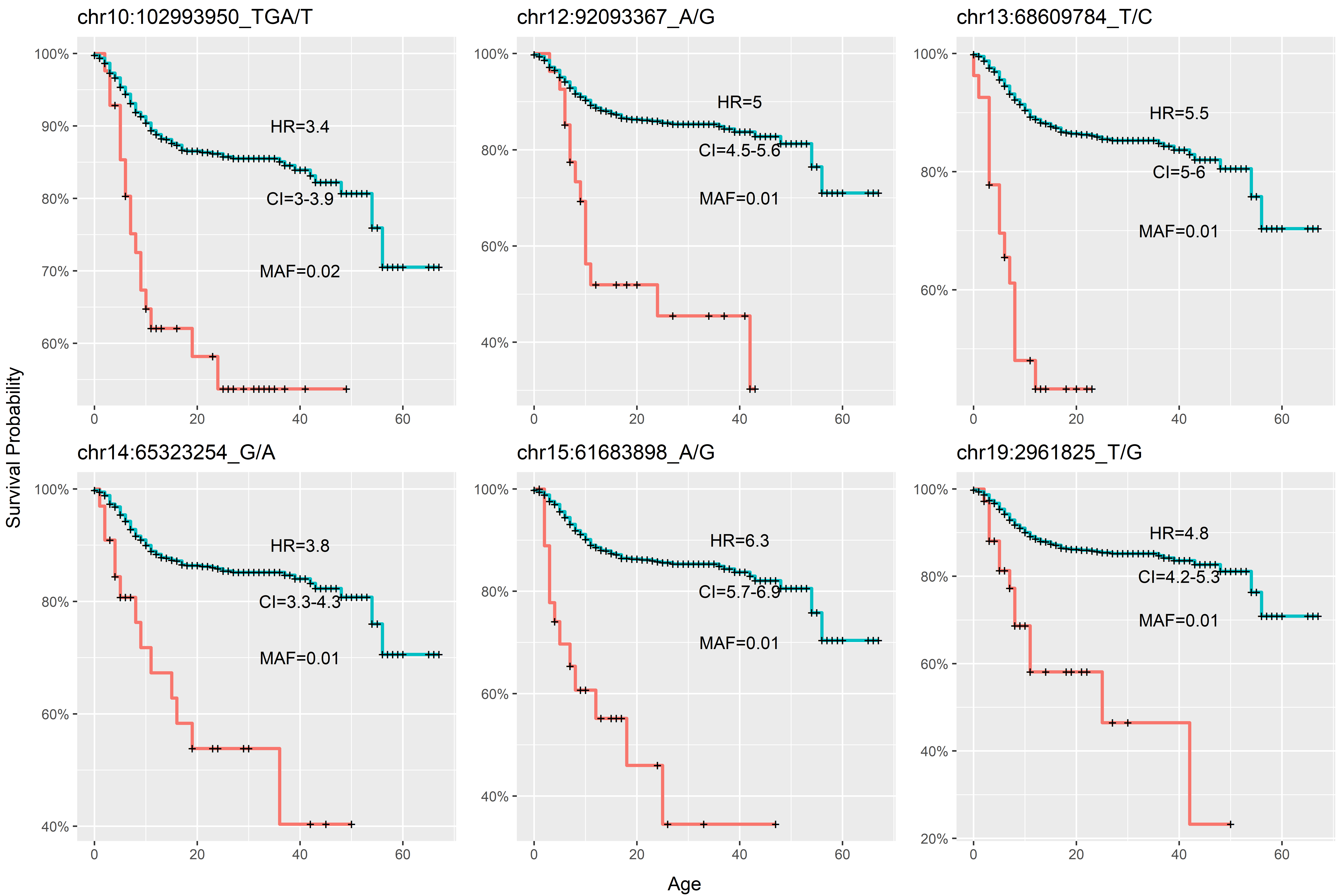

**Supplemental Figure 4.**

Kaplan-Meier plots of stroke probability for homozygous non-effect (blue), heterozygous and (if n>0) homozygous effect (light red) individuals over time in years for each candidate variant. ‘+’ represent right-censored data; HR = hazards ratio; CI = 95% confidence intervals; P = p-value from Gehan-Wilcoxon test

**Supplemental Table 2**. cis-eQTL Results

| **Chr** | **Position** | **Effect Allele** | **rsID** | **Tissue** | **Gene** | **Expression Direction** | **Posterior Probability*** |
| --- | --- | --- | --- | --- | --- | --- | --- |
| 1 | 205490782 | G | rs12144136 | Muscle - Skeletal | CDK18 | Up | -- |
| 8 | 31119876 | C | rs4733225 | Heart – Left Ventricle | WRN | Up | 0.52 |
| 8 | 31000152 | C | rs12234936 | Brain – Substantia nigra | PURG | Up | 0.50 |
| 8 | 30998948 | G | rs1362910 | Brain – Substantia nigra | PURG | Up | 0.50 |
| 12 | 91672731 | A | rs77217583 | Brain – Frontal Cortex BA9 | LUM | Down | 1 |
| 12 | 92473225 | A | rs117881990 | Brain – Hypothalamus | NUDT4 | Up | 1 |
| 14 | 64976718 | G | rs61160081 | Brain – Substantia nigra | RAB15 | Up | 1 |
| 15 | 61855008 | A | rs2241492 | Brain – Spinal cord cervical c-1 | VPS13C | Down | 1 |

*Posterior probability (PP) from Coloc analysis; rs12144136 has no PP because it was observed to be a cis-eQTL itself and was not included in the analysis.

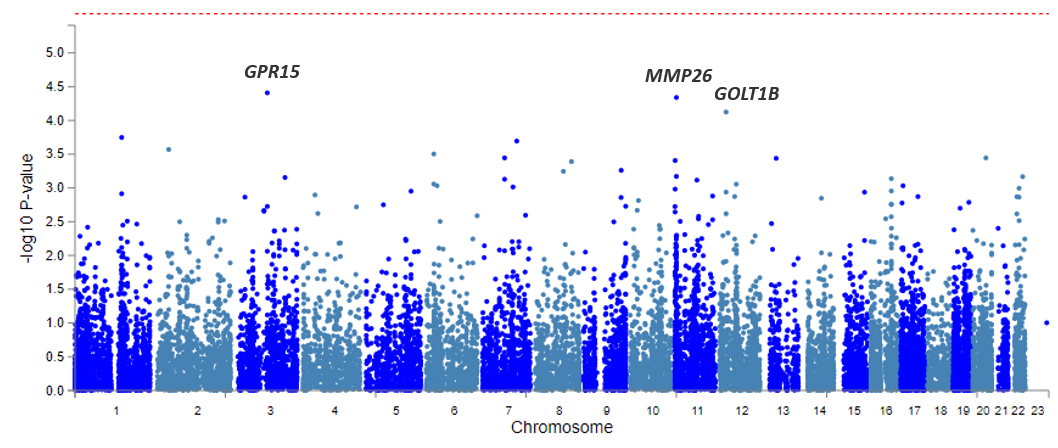

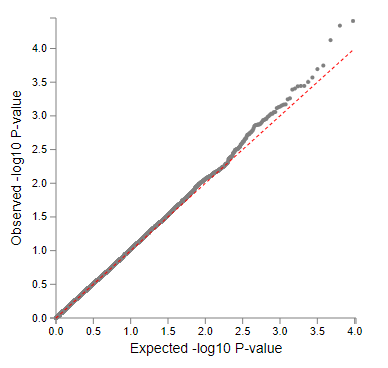

**Supplemental Figure 5.** (TOP) MAGMA gene-based association across all 14M p-values. Dotted red line represents Bonferroni-corrected alpha threshold of *P =* 2.6x10^-6^; labeled genes exhibited *P <* 1x10^-4^. (BOTTOM) qq plot of the MAGMA results.

**Supplemental Table 3.** MAGMA gene-level association testing top results.

| Ensembl Gene ID | Chr | Start | Stop | Num SNPs | Num Param | Z | P | SYMBOL |
| --- | --- | --- | --- | --- | --- | --- | --- | --- |
| ENSG00000154165 | 3 | 98250743 | 98251960 | 4 | 2 | 3.9503 | 3.90E-05 | *GPR15* |
| ENSG00000167346 | 11 | 4726157 | 5013659 | 1422 | 108 | 3.912 | 4.58E-05 | *MMP26* |
| ENSG00000111711 | 12 | 21654715 | 21671342 | 103 | 22 | 3.7904 | 7.52E-05 | *GOLT1B* |
| ENSG00000173171 | 1 | 1.55E+08 | 1.55E+08 | 7 | 3 | 3.5693 | 1.79E-04 | *MTX1* |
| ENSG00000154415 | 7 | 1.14E+08 | 1.14E+08 | 896 | 60 | 3.5371 | 2.02E-04 | *PPP1R3A* |
| ENSG00000003509 | 2 | 37458774 | 37480546 | 128 | 29 | 3.4606 | 2.69E-04 | *NDUFAF7* |
| ENSG00000146047 | 6 | 25727137 | 25727573 | 8 | 4 | 3.4192 | 3.14E-04 | *HIST1H2BA* |
| ENSG00000165171 | 7 | 73248920 | 73256865 | 32 | 6 | 3.3825 | 3.59E-04 | *WBSCR27* |
| ENSG00000166913 | 20 | 43514317 | 43537173 | 77 | 16 | 3.3821 | 3.60E-04 | *YWHAB* |
| ENSG00000150907 | 13 | 41129804 | 41240734 | 357 | 38 | 3.3784 | 3.64E-04 | *FOXO1* |
| ENSG00000070031 | 11 | 626431 | 627143 | 2 | 1 | 3.3578 | 3.93E-04 | *SCT* |
| ENSG00000155792 | 8 | 1.21E+08 | 1.21E+08 | 1057 | 63 | 3.3479 | 4.07E-04 | *DEPTOR* |
| ENSG00000171484 | 9 | 1.25E+08 | 1.25E+08 | 7 | 2 | 3.2643 | 5.49E-04 | *OR1B1* |
| ENSG00000212999 | 8 | 93895865 | 93898013 | 6 | 4 | 3.2544 | 5.68E-04 | *AC117834.1* |
| ENSG00000176937 | 11 | 4824663 | 4825847 | 6 | 2 | 3.2052 | 6.75E-04 | *OR52R1* |
| ENSG00000100304 | 22 | 43562628 | 43583139 | 188 | 47 | 3.2027 | 6.81E-04 | *TTLL12* |
| ENSG00000163661 | 3 | 1.57E+08 | 1.57E+08 | 33 | 11 | 3.1941 | 7.01E-04 | *PTX3* |
| ENSG00000188038 | 16 | 67918708 | 67922758 | 13 | 5 | 3.1831 | 7.29E-04 | *NRN1L* |
| ENSG00000189143 | 7 | 73213872 | 73247014 | 153 | 23 | 3.1771 | 7.44E-04 | *CLDN4* |
| ENSG00000054967 | 11 | 73087309 | 73108519 | 83 | 20 | 3.1682 | 7.67E-04 | *RELT* |

**Chr** = Chromosome; **Start, Stop** = b38 genomic coordinates; **Num SNPs** = SNPs observed in current GWAS; **Num Param** = number of co-variates in the MAGMA model; **Z** = probit transform of gene-level p-value from GWAS summary statistics; ***P*** *=* observed p-value from MAGMA test.

**Supplemental Table 4.** MAGMA gene-set association results (*P* < 0.001).

| Num Genes | BETA | STD | SE | P | Gene Set Name |
| --- | --- | --- | --- | --- | --- |
| 11 | 0.93609 | 0.022505 | 0.23587 | 3.63E-05 | Platelet dense tubular network |
| 10 | 0.93322 | 0.021393 | 0.27892 | 0.000411 | Adenylate cyclase_activating_dopamine_receptor_signaling_pathway |
| 19 | 0.70913 | 0.022402 | 0.21854 | 0.000589 | Gas_transport |
| 74 | 0.32628 | 0.020312 | 0.10106 | 0.000623 | Extracellular_matrix_disassembly |
| 20 | 0.5433 | 0.017608 | 0.16842 | 0.000629 | Regulation_of_double_strand_break_repair_via_nonhomologous_end_joining |
| 9 | 0.84484 | 0.018373 | 0.26332 | 0.000669 | Platelet_dense_tubular_network_membrane |
| 11 | 0.98545 | 0.023692 | 0.30811 | 0.000692 | Hemoglobin_complex |
| 36 | 0.45434 | 0.019748 | 0.14324 | 0.000759 | Regulation_of_systemic_arterial_blood_pressure_by_hormone |
| 17 | 0.54179 | 0.01619 | 0.17193 | 0.000814 | Regulation_of_translational_fidelity |
| 7 | 1.1608 | 0.022265 | 0.36929 | 0.000836 | Regulation_of_retina_development_in_camera_type_eye |
| 96 | 0.27511 | 0.019496 | 0.087549 | 0.000839 | Alpha_beta_t_cell_differentiation |
| 5 | 1.317 | 0.02135 | 0.42535 | 0.000982 | Female_germ_cell_nucleus |

**Num Genes** = number of genes in the gene set; **BETA** = MAGMA regression coefficient; **STD** = standard deviation; **SE** = standard error

**Supplemental Table 5**

| **Chr** | **Gene** | **Evidence** | **P-valuesⴕ** | **Gene Description** | **Previous Studies Linking Gene to Stroke or Other Cardiovascular Disease and Risk Factors** |
| --- | --- | --- | --- | --- | --- |
| 1 | *CDK18* | proximity & cis-eQTL | 2.38 x 10^-9^ | Kinase involved in cell division | CDK18 variants associated with early-onset ischemic stroke in non-SCD Japanese cohort (Yamada et al., 2017) |
| 2 | *SH3RF3* | proximity | 3.35 x 10^-8^ | Ubiquitin protein ligase and positive regulator of JNK cascade | Variants in gene region linked to coronary heart disease (https://pubmed.ncbi.nlm.nih.gov/28396569/). |
| 5 | *ADAMTS2* | proximity | 3.70 x 10^-9^ | Extracellular collagen assembly | Intronic variants within gene ADAMTS2 associated with pediatric stroke (Arning et al., 2012) |
| 6 | *ERMARD* | proximity | 2.52 x 10^-9^ | Endoplasmic reticulum RNA degradation | -- |
| 7 | *PKD1L1* | proximity | 2.71 x 10^-8^ | Ciliary calcium channel regulation | Variants in PKD1L1 linked to congenital heart disease (Vetrini et al., 2016), gene involved in lung & heart lateralization in mice (Grimes et al., 2016) |
| 7 | *CUX1* | proximity | 3.70 x 10^-9^ | Homeodomain-containing transcription factor | GWAS Catalog – mean platelet volume, reticulocyte count |
| 8 | *WRN* | coloc cis-eQTL | 3.10 x 10^-8^ | RecQ like DNA helicase | GWAS catalog – cardiovascular disease, HDL cholesterol |
| 10 | *KIAA1217* | proximity | 4.43 x 10^-8^ | Unknown | GWAS catalog – coronary artery disease, mean corpuscular hemoglobin, reticulocyte volume |
| 10 | *CNNM2* | proximity | 3.90 x 10^-8^ | Cation transport mediator | *CNNM2* gene expression associated with heart attack (https://www.ncbi.nlm.nih.gov/pmc/articles/PMC5027110/) |
| 10 | *NT5C2* | proximity | 3.90 x 10^-8^ | Hydrolase; maintains purine levels | GWAS Catalog – Cardiovascular disease (rs11191580), coronary artery disease (rs11191559), and other cardiovascular disease markers |
| 12 | *BTG1* | proximity | 2.51 x 10^-8^ | Antiproliferation | Up-regulated in stroke-associated carotid plaques (https://www.ncbi.nlm.nih.gov/pmc/articles/PMC3170468/) |
| 12 | *LUM* | coloc cis-eQTL | NA | Extracellular matrix-localized peptidoglycan | Inflammation-associated protein upregulated in serum after aortic surgery (https://www.ncbi.nlm.nih.gov/pmc/articles/PMC7932520/) |
| 12 | *NUDT4* | coloc cis-eQTL | NA | Nudix hydrolase | GWAS Catalog – mean corpuscular hemoglobin, reticulocyte count |
| 13 | *RPS3AP52* | proximity | 3.78 x 10^-12^ | Ribosomal Protein S3a Pseudogene | -- |
| 14 | *FUT8* | proximity | 2.58 x 10^-9^ | Fucosyltransferase | Ischemic stroke (https://pubmed.ncbi.nlm.nih.gov/32474671/) |
| 14 | *RAB15* | coloc cis-eQTL | NA | Ras-related Protein | -- |
| 15 | *VPS13C* | proximity & cis-eQTL | 5.40 x 10^-10^ | Vacuolar Protein Sorting-Associated Protein | -- |
| 19 | *TLE6* | proximity | 2.25 x 10^-8^ | Groucho/ transducin-like Enhancer | -- |

**Supplemental Table 6.** Enrichment test results using the GTEX v8 database of tissue specific gene expression of 18 candidate genes. Shown are tissues with *P* < 0.05 (FDR).

| **Category** | **Tissue Type** | **Num Genes** | **Observed** | **FDR** | **Gene IDs** |
| --- | --- | --- | --- | --- | --- |
| One-sided test – UP | Artery Coronary | 2038 | 8 | 0.000464 | ENSG00000117266:ENSG00000172985:ENSG00000087116: |
|  |  |  |  |  | ENSG00000257923:ENSG00000148842:ENSG00000139329: |
|  |  |  |  |  | ENSG00000173598:ENSG00000139998 |
| Two-sided test | Brain Hypothalamus | 8435 | 13 | 0.005112 | ENSG00000117266:ENSG00000172985:ENSG00000087116: |
|  |  |  |  |  | ENSG00000257923:ENSG00000165392:ENSG00000120549: |
|  |  |  |  |  | ENSG00000076685:ENSG00000133639:ENSG00000139329: |
|  |  |  |  |  | ENSG00000173598:ENSG00000139998:ENSG00000129003: |
|  |  |  |  |  | ENSG00000104953 |
| One-sided test – DOWN | Spleen | 2069 | 7 | 0.005371 | ENSG00000172985:ENSG00000257923: |
|  |  |  |  |  | ENSG00000120549:ENSG00000148842: |
|  |  |  |  |  | ENSG00000139329:ENSG00000173598:ENSG00000139998 |
| Two-sided test | Artery Coronary | 2987 | 8 | 0.007412 | ENSG00000117266:ENSG00000172985:ENSG00000087116: |
|  |  |  |  |  | ENSG00000257923:ENSG00000148842:ENSG00000139329 |
|  |  |  |  |  | :ENSG00000173598:ENSG00000139998 |
| One-sided test -DOWN | Pancreas | 9586 | 13 | 0.021339 | ENSG00000117266:ENSG00000172985:ENSG00000087116: |
|  |  |  |  |  | ENSG00000130023:ENSG00000257923:ENSG00000165392: |
|  |  |  |  |  | ENSG00000120549:ENSG00000148842:ENSG00000076685: |
|  |  |  |  |  | ENSG00000173598:ENSG00000033170:ENSG00000139998: |
|  |  |  |  |  | ENSG00000129003 |
| One-sided test -DOWN | Esophagus Mucosa | 3580 | 8 | 0.02624 | ENSG00000117266:ENSG00000172985:ENSG00000087116: |
|  |  |  |  |  | ENSG00000148842:ENSG00000173598:ENSG00000033170: |
|  |  |  |  |  | ENSG00000139998:ENSG00000129003 |
| Two-sided test | Esophagus Mucosa | 5811 | 10 | 0.029758 | ENSG00000117266:ENSG00000172985:ENSG00000087116: |
|  |  |  |  |  | ENSG00000120549:ENSG00000148842:ENSG00000076685: |
|  |  |  |  |  | ENSG00000173598:ENSG00000033170:ENSG00000139998: |
|  |  |  |  |  | ENSG00000129003 |
| One-sided test – UP | Artery Aorta | 2756 | 7 | 0.031929 | ENSG00000117266:ENSG00000172985:ENSG00000257923: |
|  |  |  |  |  | ENSG00000148842:ENSG00000076685:ENSG00000139329: |
|  |  |  |  |  | ENSG00000173598 |
| Two-sided test | Artery Tibial | 4738 | 9 | 0.032903 | ENSG00000117266:ENSG00000172985:ENSG00000120549: |
|  |  |  |  |  | ENSG00000148842:ENSG00000076685:ENSG00000139329: |
|  |  |  |  |  | ENSG00000173598:ENSG00000033170:ENSG00000139998 |
| One-sided test -DOWN | Brain Hypothalamus | 5904 | 10 | 0.033985 | ENSG00000172985:ENSG00000087116:ENSG00000257923: |
|  |  |  |  |  | ENSG00000165392:ENSG00000120549:ENSG00000076685: |
|  |  |  |  |  | ENSG00000133639:ENSG00000139329:ENSG00000173598: |
|  |  |  |  |  | ENSG00000129003 |
| One-sided test -DOWN | Brain Substantia nigra | 7162 | 11 | 0.03424 | ENSG00000172985:ENSG00000087116:ENSG00000130023: |
|  |  |  |  |  | ENSG00000257923:ENSG00000165392:ENSG00000120549: |
|  |  |  |  |  | ENSG00000076685:ENSG00000133639:ENSG00000139329: |
|  |  |  |  |  | ENSG00000173598:ENSG00000129003 |
| Two-sided test | Brain Substantia nigra | 8807 | 12 | 0.047278 | ENSG00000117266:ENSG00000172985:ENSG00000087116: |
|  |  |  |  |  | ENSG00000130023:ENSG00000257923:ENSG00000165392: |
|  |  |  |  |  | ENSG00000120549:ENSG00000076685:ENSG00000133639: |
|  |  |  |  |  | ENSG00000139329:ENSG00000173598:ENSG00000129003 |
| Two-sided test | Pancreas | 10331 | 13 | 0.048235 | ENSG00000117266:ENSG00000172985:ENSG00000087116: |
|  |  |  |  |  | ENSG00000130023:ENSG00000257923:ENSG00000165392: |
|  |  |  |  |  | ENSG00000120549:ENSG00000148842:ENSG00000076685: |
|  |  |  |  |  | ENSG00000173598:ENSG00000033170:ENSG00000139998: |
|  |  |  |  |  | ENSG00000129003 |

**Category** = GTEx defined differential gene expression category; **Num genes**= number of background genes in that gene-set with average expression > 1; **Num observed** = number of genes used in the hypergeometric enrichment test; **FDR** = false discovery rate adjusted p-value; **Genes** = Ensembl gene IDs for the genes used in the enrichment test.

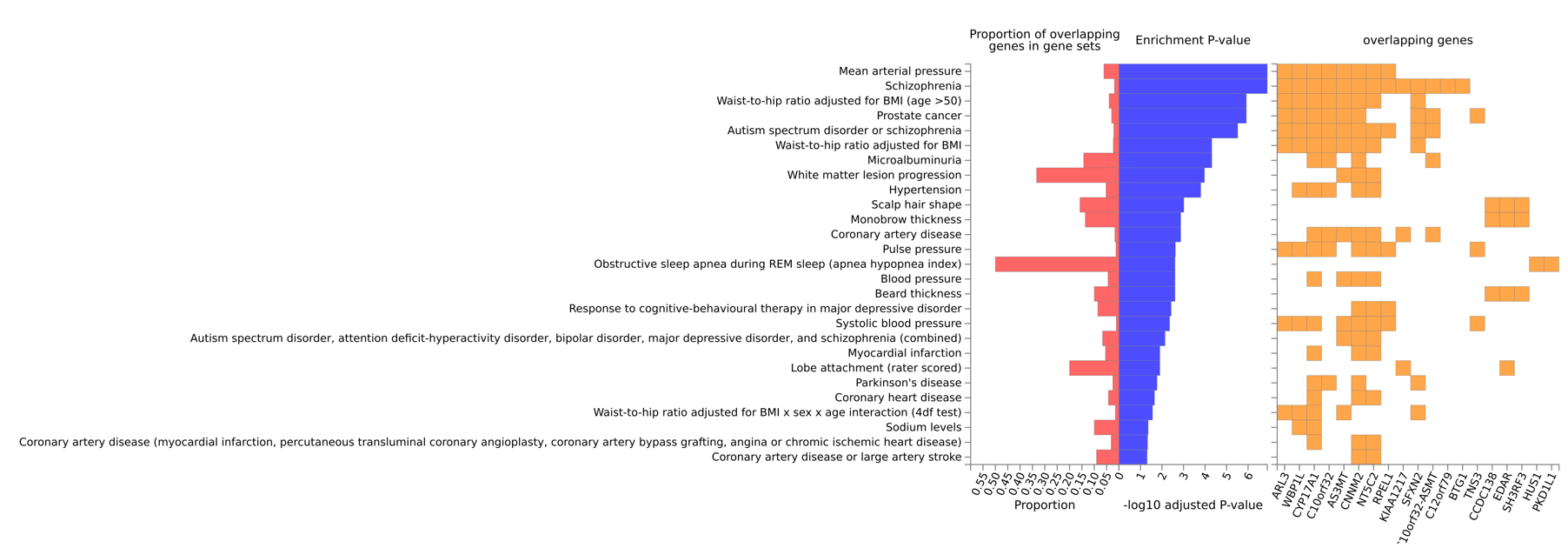

**Supplemental Figure 6. Gene set enrichment of 35 candidate genes against GWAS Catalog.** Genes from the GWAS results were selected based on location within 100kb of one of the 14 lead SNPs. In addition, the targets of cis-eQTLs which are located within 10kb of a lead SNP were also included (*WRN, LUM, NUDT4*). Red, proportion of genes; blue, transformed p-values (FDR); yellow, genes.

**Supplemental Table 7.** Candidate SNPs for stroke risk in SCD individuals from literature (left), as well as result from the current study (right).

| **Literature** | | | | | | | **Brazil SCD** | | | | |
| --- | --- | --- | --- | --- | --- | --- | --- | --- | --- | --- | --- |
| SNP ID | **Gene** | **Position** | **Effect Allele** | **Reference*** | **Published Effect** | **Study Type** | **Effect AF (%)** | **HR (95% CI)** | **P-value** | **Lowest P-val in 1Mb region** | **Site with lowest P-val in 1Mb region** |
| rs1041163 | VCAM1 | 1:100718269 | C | 1 | Risk | Candidate | 23.6 | 0.86 (0.6-1.12) | 0.26 | 1.70E-03 | 1:100515235:C:G |
| rs3783613 | VCAM1 | 1:100731231 | C | 2 | Protective | Candidate | 7.1 | 1.44 (1.07-1.8) | 0.05 | 1.70E-03 | 1:100515235:C:G |
| rs284875 | TGFBR3 | 1:91705151 | T | 3,4,5 | Risk | Candidate | 7.4 | 0.97 (0.65-1.44) | 0.87 | 1.50E-03 | 1:91652079:C:T |
| rs3732410 | GOLGB1 | 3:121696873 | C | 6 | Protective | GWAS | 14.0 | 0.78 (0.56-1.09) | 0.14 | 1.19E-07 | 3:122057039:GT:G |
| rs1042714 | ADRB2 | 5:148826910 | G | 1 | Protective | Candidate | 22.6 | 1.43 (1.08-1.89) | 0.01 | 3.40E-05 | 5:149164128:C:G |
| rs730012 | LTC4S | 5:179793637 | C | 7 | Protective | Candidate | 13.9 | 1.06 (0.78-1.42) | 0.73 | 3.70E-09 | 5:179375279:G:A |
| rs1044498 | ENPP1 | 6:131851228 | C | 6,5,8 | Conflicting | Both | 48.8 | 1.05 (0.84-1.30) | 0.68 | 4.05E-05 | 6:131718438:C:T |
| rs1800629 | TNF-a | 6:31575254 | A | 1,7,8,5 | Conflicting | Candidate | 11.5 | 0.97 (0.69-1.37) | 0.88 | 6.38E-05 | 6:31334053:C:T |
| rs489347 | TEK | 9:27187218 | G | 3,4,5 | Risk | Candidate | 27.5 | 1.10 (0.86-1.41) | 0.45 | 4.01E-05 | 9:27338186:T:C |
| rs11853426 | ANXA2 | 15:60317231 | T | 3,4 | Risk | Candidate | 45 | 0.91 (0.74-1.13) | 0.39 | 2.66E-04 | 15:60717990:G:C |
| rs1805015 | IL4R | 16:27362859 | C | 1,7 | Risk | Candidate | 28.5 | 0.98 (0.77-1.24) | 0.84 | 2.62E-06 | 16:27857484:G:A |
| rs2238432 | ADCY9 | 16:3964140 | A | 3,4 | Protective | Candidate | 17.3 | 0.98 (0.74-1.30) | 0.88 | 2.67E-04 | 16:4158065:GA:G |
| rs5742911 | LDLR | 19:11132769 | G | 1 | Protective | Candidate | 25.6 | 1.09 (0.86-1.38) | 0.46 | 1.54E-03 | 19:11091840:G:C |

**1** = Hoppe et al 2004, Blood; **2** = Taylor et al 2002, Blood; **3** = Sebastiani et al 2005, Nat Gen; **4** = Flanagan et al 2011, Blood; **5** = Belisario et al 2016 Ann Hem; **6** = Flanagan et al 2013, Blood; **7** = Hoppe et al 2007, Stroke; **8** = Belisario et al 2015, Blood. **Study type** = candidate gene, GWAS, or both; **AF** = allele frequency; **HR** = hazard ratio; **CI** = confidence interval; **P-value** = observed p-value for the candidate SNP in current study.

**Supplemental Table 8.** Overlap between current SCD ischemic stroke GWAS and previous studies in non-SCD early onset ischemic stroke GWAS.

| **Literature** | | | | | | | | **Brazil SCD** | | | | | | |
| --- | --- | --- | --- | --- | --- | --- | --- | --- | --- | --- | --- | --- | --- | --- |
| **SNP ID** | **Gene** | **Position** | **Effect Allele** | **Published OR** | **Phenotype** | **Ancestry Group** | **Sample Size** | **Effect Allele** | **Effect AF (%)** | **HR** | **P-value** | **Site with lowest P-val in 500kb region*** | **HR**  **(95% CI)** | **Lowest P-val in 500kb region** |
| rs77571454 | *CDK18* | 1:205,531,350 | A | 1.72 | Early onset ischemic stroke | East Asian | 6,224 (450 cases) | Not Observed | -- | -- | -- | 1:205490782:T:G | 2.7  (2.37 - 3.3) | 2.38x10^-9^ |
| rs1364044 | *ADAMTS12* | 5:33,879,687 | T | 1.84 | Pediatric stroke, ≤18 | European | 270 trios | T | 40.7 | 1.07 | 0.5 | 5:34245124:C:T | 5.24  (4.47 – 6.01) | 2.43x10^-5^ |
| rs469568 | *ADAMTS2* | 5:179,236,407 | A | 1.87 | Pediatric stroke, ≤18 | European | 270 trios | A | 55.4 | 1.14 | 0.22 | 5:179375279:G:A | 5.36 (4.81 – 5.91) | 3.70x10^-9^ |
| rs11196288 | HABP2 | 10:113,297,684 | G | 1.41 | Young onset stroke | European, South Asian, African American | 25,594 (3,626 cases) | G | 5.5 | 1.4 | 0.16 | 10:112897510:TTA:T | 2.72 (4.07 – 3.37) | 2.58x10^-3^ |

**OR** = odds ratio; **AF** = allele frequency; **HR** = hazard ratio; **P-value** = observed p-value for the candidate SNP in current study; **CI** = confidence interval.
